# Estimation and Interactive Visualization of the Time-Varying Reproduction Number *R*_*t*_ and the Time-Delay from Infection to Estimation

**DOI:** 10.1101/2020.09.19.20197970

**Authors:** Fabian Valka, Carla Schuler

## Abstract

*R*_*t*_ plays a key role in the development of the COVID-19 pandemic. The methods used for building an interactive website for the visualization of the time-varying reproduction number *R*_*t*_ and a novel way to visualize the time delay from infection to estimation overlayed with the *R*_*t*_ estimate are described and analyzed with regards to the influence of parameters chosen and compared to published estimates for Austria. Visualizing the time delays enables the interactive exploration of suspected changes in transmission and their effect on the *R*_*t*_ estimate.

## 1 Introduction

An interactive web-based tool was developed for visualizing and exploring the time-varying reproduction number and time delays from infection to onset, to reporting and inclusion in the estimation of *R*_*t,τ*_ on a regional level for Austria. Users can choose the Austrian state, sliding time window length *τ* and overlay the AGES *R*_*t*,13_ estimate [1].

The tool is available as free open-source software on GitHub: https://github.com/fvalka/r_estimate.

### 1.1 Importance of Time Delays from a Control Theory Perspective

The introduction, discontinuation, reintroduction, and change of non-pharmaceutical interventions can also be viewed from a control theory perspective.[2] On-off style control of introduction and lifting of lockdown measures throughout the post-pandemic period has been explored.[3]

Introducing time-delays into otherwise stable closed control-loops can make them unstable.[4, 5] Careful treatment and consideration of the time-delays involved in the available observables is therefore essential.

## 2 Estimation of the Time-Varying Reproduction Number *R*_*t*_

In analogy to the method published by Richter, et al.[6] from AGES, the time-varying reproduction number *R*_*t*_ is estimated according to Cori, et al.[7] Sliding time windows of length *τ* days are used with the assumption that within the time window *τ* the reproduction number *R*_*t*_ is constant, defining *R*_*t,τ*_.

The R-package EpiEstim[8] provided by Cori, et al. is used for the implementation.

### 2.1 Serial Interval

Accuracy of the serial interval estimation plays a key role in the accuracy of the estimation of *R*_*t,τ*_.

Representing the uncertainties contained in the current serial interval estimates[9] was realized using the uncertain_si method in estimate_R. This samples the parameters for the serial intervals Gamma distribution Gamma_*s*_ from a truncated normal distribution. The median of the *R*_*t,τ*_ estimate is only affected to a limited extent by this, but the credible interval will be increased around those median values[7], giving a more accurate representation based on our current beliefs about the serial interval.

Parameters used in the implementation are documented in Table 1.

**Table 1:**
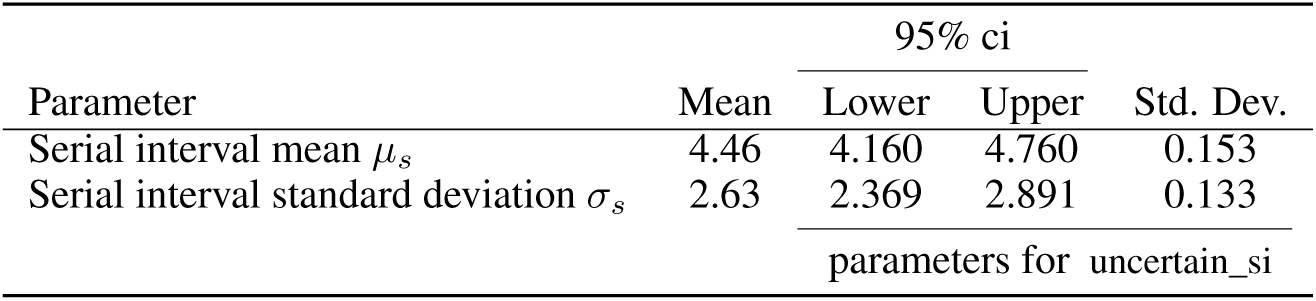
Serial interval Gamma distribution Gamma_*s*_ parameters based upon estimates by Richter, et al. for Austria.[9]

Standard deviations of the mean serial interval and the standard deviation of the standard deviation were obtained based upon the assumption of normality from the 95% confidence interval CI_*lower*_, see Equation 1.

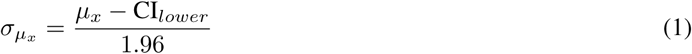

Based upon these parameters multiple serial intervals are explored by estimate_R.[8] Explored serial interval distributions are shown in Figure 3. During each estimation 500 discretized Gamma distributions of the serial interval are explored.

### 2.2 Sliding Time Window *τ*

Choosing *τ* is a trade-off between less noise and less delay. Smaller time windows *τ* lead to a more rapid detection of changes in *R*_*t,τ*_, but also to more statistical noise.[10]

Cori, et al. describe a method for choosing the time window size *τ* based upon the minimum number of cases included in each time window [10]. Based upon a chosen target coefficient of variation for the *R*_*t,τ*_ estimate of at least 0.4 a minium of 7 cases in each time window is required. With this in consideration and the usage of a time window of 7 days in Cori, et al.[7] a time window size *τ* of 7 days for the interactive visualization was chosen. AGES uses a 13 days time window for their estimation, no explanation is given for this choice and this *τ* was therefore only used for comparisons but can be chosen by the user on the interactive front-end.

The effect a change in *τ* has on the estimation of *R*_*t,τ*_ is illustrated in Figure 2. Comparing the first date where the median of *τ* has fallen below 1 illustrates the trade-off discussed above. For *τ* = 7 the condition *R*_*t*,7_ *<* 1 is first met on the 31st of March 2020, for *τ* = 13 the condition *R*_*t*,13_ *<* 1 is only met 4 days later on the 4th of April 2020.

**Figure 1:**
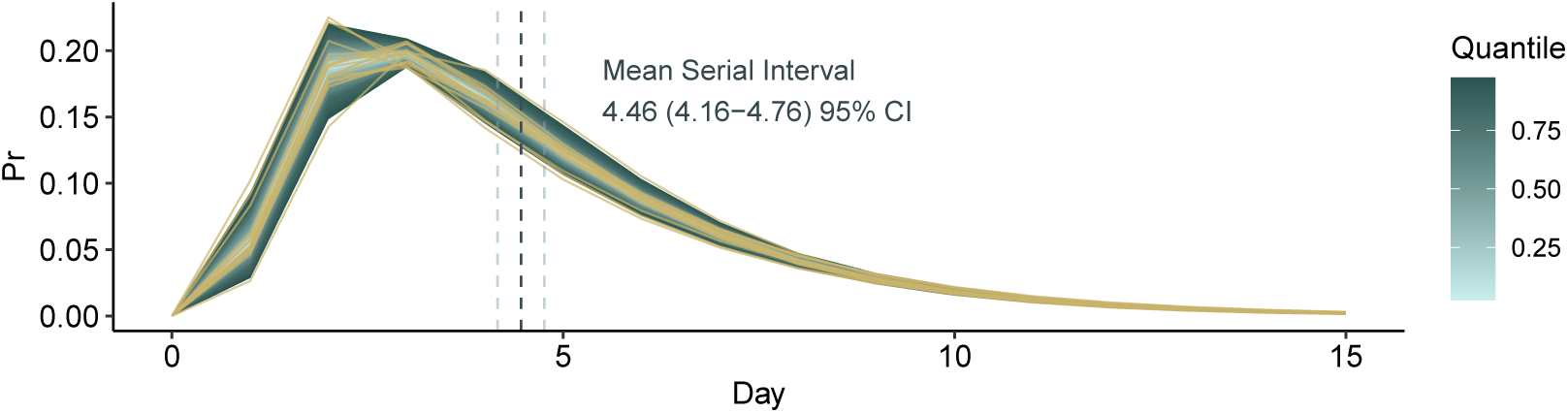
Serial interval distributions explored during *R*_*t,τ*_ estimation. Quantiles shown in color scaling (blue), with the outer border marking the 0.95 credible interval (0.025 and 0.975 quantiles). Overlayed with samples from the explored discrete serial interval distributions (yellow).

**Figure 2:**
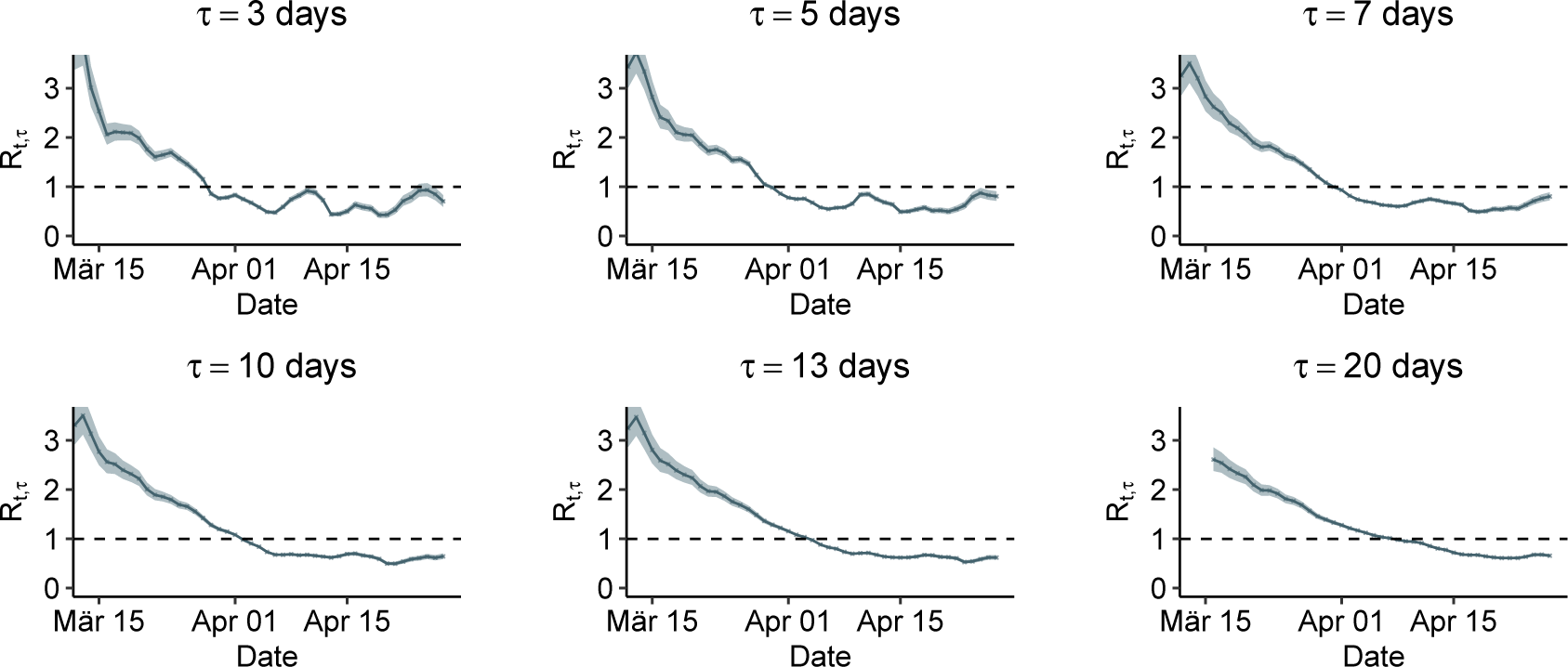
Effect of different time window, *τ*, choices on the *R*_*t,τ*_ estimate for Austria, as of 2020-04-27. Left censored at first entry in data-set *t*_0_ + *τ* Data source: data.gv.at.[11]

**Figure 3:**
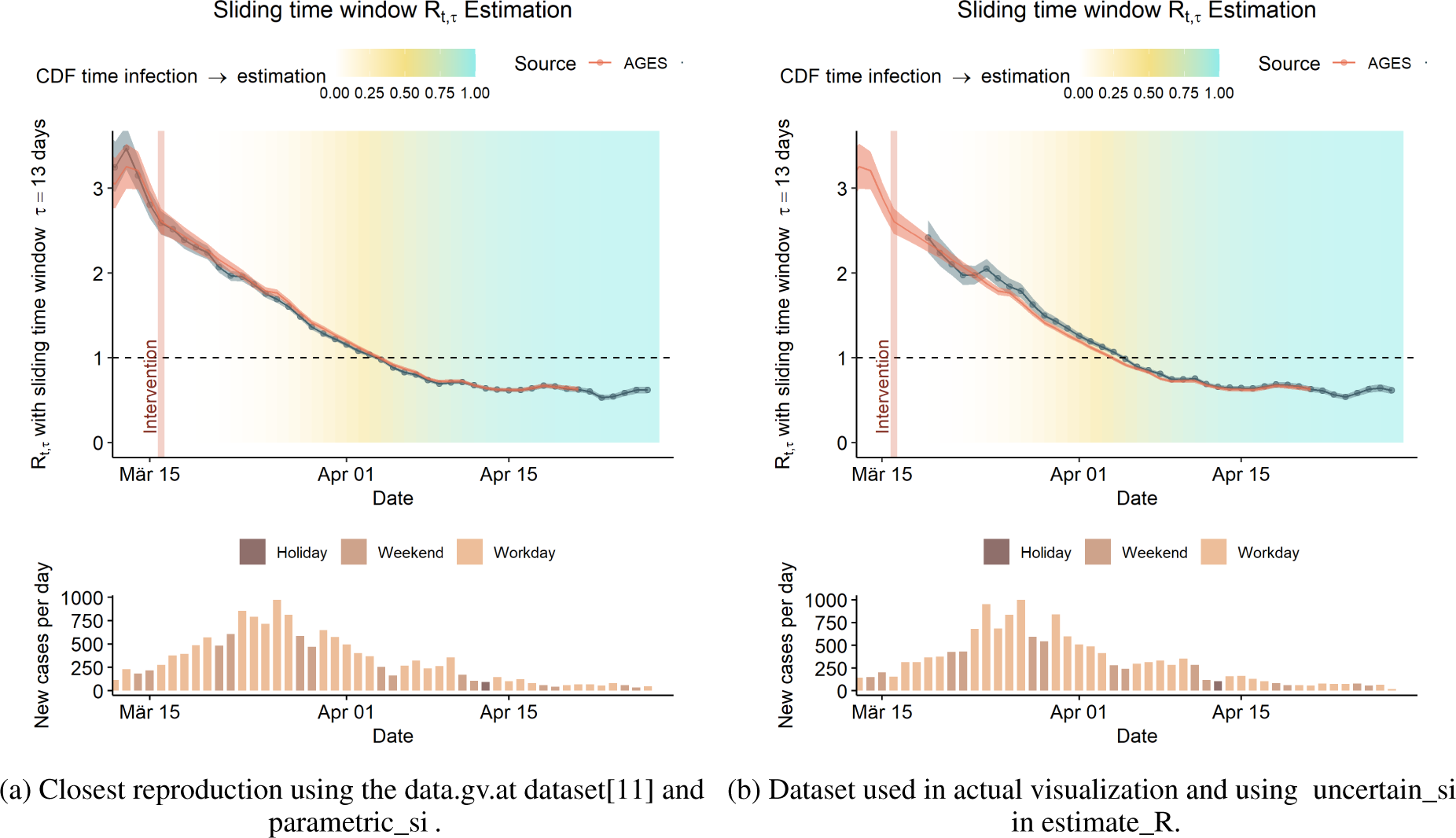
Comparison between own estimates and *R*_*t,τ*_ estimates published by AGES.[12]

**Figure 4:**
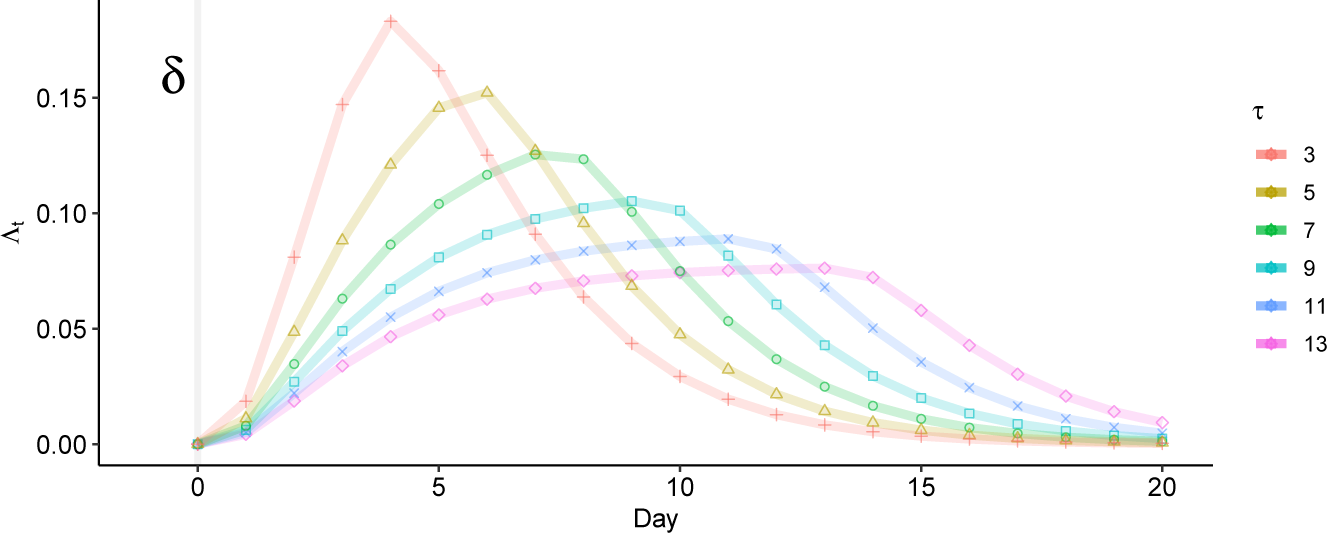
Λ_*t*_ response. Normalized such that 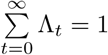

**Figure 5:**
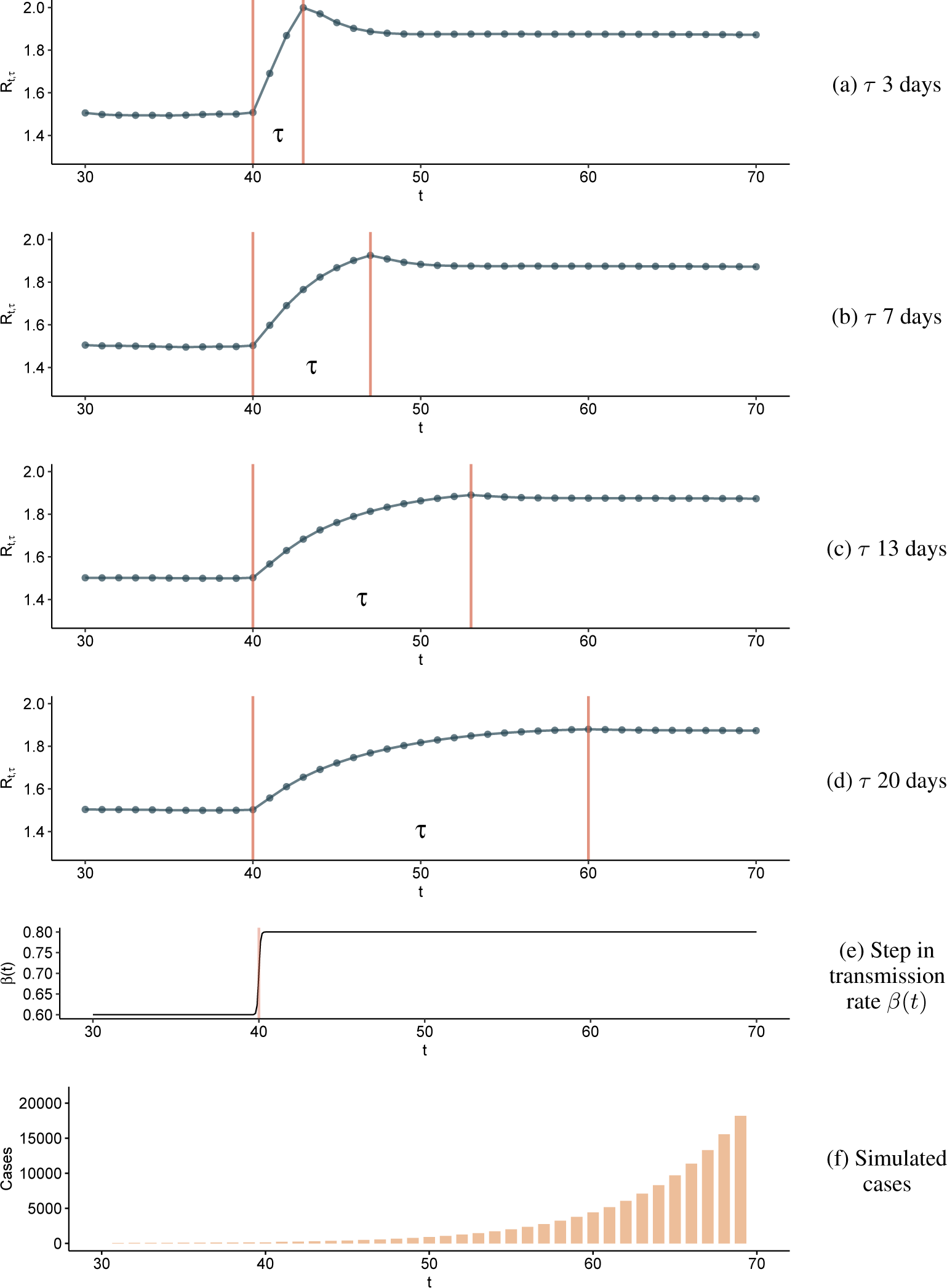
Response of the *R*_*t,τ*_ estimation to a step function in the transmission rate *β*(*t*), and therefore also in *R*_*t*_ of a SEIR model.

A 7-day window has the added advantage that it always contains exactly 2 weekend days.

### 2.3 Crossvalidation with AGES estimates for Austria

The output of the estimation described in this section was compared to the *R*_*t,τ*_ estimates obtained by AGES on the 24th of April 2020[1].

Two different methods were chosen for the comparison, one where the dataset which allows the closest reproduction of the estimates obtained by AGES was used as shown in figure Figure 3a. In the actual interactive visualization a different dataset is used. The comparison against this dataset is shown in Figure 3b. For a further discussion on the datasets see section 4.

## 3 Estimation of the time delay from infection to inclusion in the *R*_*t,τ*_ estimation

The total time delay from infection to inclusion in the estimate *t*_*infection,estimation*_ will be split up into three different time delays. The time delay from infection to onset *t*_*infection,onset*_, the incubation period. The time delay from onset to reporting *t*_*onset,reporting*_ and the time delay from reporting to inclusion in the estimation *t*_*reporting,estimation*_, which is caused by the *R*_*t,τ*_ estimation method of assuming a constant *R*_*t*_ within the time window *τ*.

### 3.1 Time delay from infection to reporting *t*_*infection,reporting*_

Both *t*_*infection,onset*_ and *t*_*onset,reporting*_ are assumed to be independent random variables and a Gamma distribution was used for the estimation. Estimates of the Gamma distribution parameters for *t*_*infection,onset*_ were taken from the supplementary appendix of the study by Zhang, et al.[13] based upon case data from Mainland China, excluding Hubei province. The parameters used are *α*_*infection,onset*_ = 4.23(SD 1.28) and *β*_*infection,onset*_ = 0.81(SD 0.24) with a mean of 5.2days and a 95% CI of the mean of (95% CI: 1.5 − 11.3) days.

Onset to reporting Gamma distribution parameters were also taken from the estimates from Mainland China, excluding Hubei[13] from the second period (Jan 28 – Feb 17) were used. Those parameters were obtained from 2079 observations and are *α*_*onset,reporting*_ = 3.18 and *β*_*onset,reporting*_ = 0.59 giving a mean of 5.3 (95% CI: 1.2 − 13.1) days.

In order to obtain the empirical CDF of *t*_*infection,reporting*_ random samples were drawn from the Gamma distributions of *t*_*infection,onset*_ and *t*_*onset,reporting*_ using the R-function **rgamma**[14]. Random samples were added up pair-wise as shown in Equation 2.

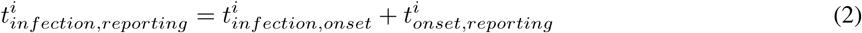

A total of 10^7^ samples were drawn from each distribution and the resulting emperical distribution function 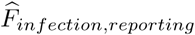 was obtained by applying the ecdf function[14] to the list of.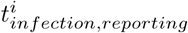

### 3.2 Timedelay from reporting to inclusion in the *R*_*t,τ*_ estimation *t*_*reporting,estimation*_

To derive the time-delay from reporting to estimation *t*_*reporting,estimation*_ we need to consider the method used for the *R*_*t,τ*_ estimation in detail. The mean of *R*_*t,τ*_ given the parameters *a* and *b* of the Gamma distribution prior is given by Equations 3 and 4.[10]

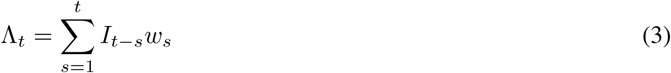

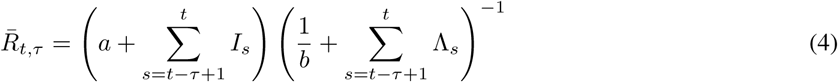

An initial investigation of the time delay was performed by investigating the propagation of a time pulse through Λ_*t*_, the key component of the denominator, as defined in defined in Equation 3. This time pulse was modeled as a dirac *δ* function at *t* = 0. The estimation process is discretized such that *t* ∈ ℕ, therefore, the discrete number of cases for the dirac *δ* is given in Equation 5.

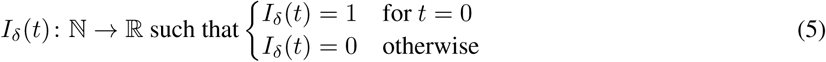

#### 3.2.1 Simulation of the Estimation Driven Time-Delay using a Compartmental Model

The *R*_*t,τ*_ response to a step function in the transmission rate *β* of a SEIR model[15] has been used for deriving the *τ* dependent time-delay inherent to the *R*_*t,τ*_ estimation method.

Parameters of the model itself are not related to COVID-19 and the model has been defined without vital dynamics.

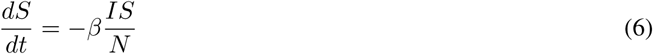

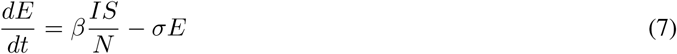

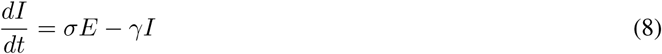

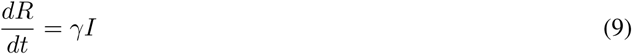

*S, E, I*, and *R* represent the total number of susceptible, exposed, infected, and removed individuals in a total population *N*, respectively. *β* denotes the transmission rate, *γ* is the removale rate, *σ* is the rate at which individuals transition from the exposed class to the infectious class. The dynamics of the model are defined in equations 6, 7, 8 and 9.[15]

For simulating a change in transmission the transmission rate *β* has been given a time dependency *β*(*t*) with a step function, modeled using a logistic function to keep the model continuously differentiable. The model can change between two different transmission rates *β*_0_, before *t*_*swap*_ and *β*_1_ after *t*_*swap*_. The steepness of the curve, *k*, was set to 20.

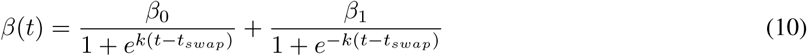

Considering the relationship between *β*(*t*) and *R*_0_ of the SEIR model, Equation 11[15], a change in *β* is proportional to a change in *R*_0,SEIR_(*t*).

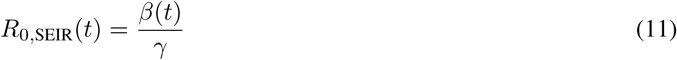

Since the dynamics of the *R*_*t,τ*_ response to a step function depends on the serial interval distribution, as can be seen in Equation 3, the actual serial interval distribution, as described in subsection 2.1, was used. For this simulation the parametric_si mode was used using the mean of the serial interval mean and the mean of the serial interval standard deviation as shown in Table 1.

In the SEIR model the average time in the infectious compartment *γ*^*−*1^ was set to 3. The average time in the exposed compartment, *σ*^*−*1^, was set to 4, the transmission rate before the step *β*_0_ was set at 0.6, the center of the logistic function, in Equation 10, *t*_*swap*_ was set to 40 and the transmission rate transitions to *β*_1_ around this point, which was set to 0.8.

For each *τ* a step response *R*_*t,τ*,sim_ is obtained from the simulation. This step response is normalized such that it is at 0before the time step is applied and that the last element *R*_*τ,τ*,sim_ = 1.

The resulting time responses from the simulations, *R*_*t,τ*,sim_, are then applied to the empirical distribution function 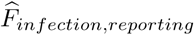 to obtain the empirical distribution function of the total time delay 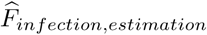 Each element of this function is calculated using Equation 12, using the definition *R*_0,*τ*,sim_ = 0.

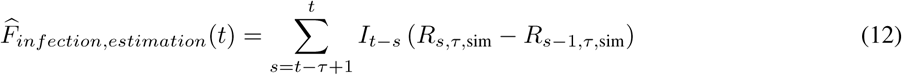

## 4 Data Sources

Case data for Austria are obtained from a GitHub repository maintained by the Complexity Science Hub Vienna based upon the official data published by the Sozialministerium. This dataset contains the case numbers for the 3 pm CEST closing time.

### 4.1 Data Correction

Some days in the Austrian states contain a negative number of cases on a given day. Those negative numbers are corrected by adding the negative correction to the previous day and adjusting the day containing the negative number to 0, keeping the total sum of cases constant.

### 4.2 Data Quality Issues

Data published by the Austrian Sozialministerium combines data into bins based upon different dates, depending on the availability of the information for each given case.[16] Currently cases are assigned to a bin by the date in the following order: Symptom onset, laboratory confirmation, reporting. This could potentially have a big impact on the estimation of *R*_*t,τ*_ and also on the time-delay involved. It is currently unknown how many of the data entries binned using which date. Estimates of *R*_*t,τ*_ published by AGES[1] therefore differ from the reproduction of their method. Comparing the results of the estimation using this dataset and the AGES estimation suggests that there might be a limited difference between those datasets, see Figure 3a.

## Data Availability

The source code, model and distribution parameters are available as open source online.
Austrian COVID-19 reported cases used in the reprint are available in the Austrian open data portal.

https://github.com/fvalka/r_estimate

https://www.data.gv.at/covid-19/

## References

[1] Lukas Richter, Daniela Schmid, Ali Chakeri, Sabine Maritschnik, Sabine Pfeiffer, and Ernst Stadlober. Epidemiologische Parameter des COVID19 Ausbruchs - Update 24.04.2020, Österreich, 2020. 2020.

[2] Greg Stewart, Klaske van Heusden, and Guy A. Dumont. How Control Theory Can Help Us Control COVID-19 - IEEE Spectrum, May 2020. Library Catalog: spectrum.ieee.org.

[3] Stephen M. Kissler, Christine Tedijanto, Edward Goldstein, Yonatan H. Grad, and Marc Lipsitch. Projecting the transmission dynamics of SARS-CoV-2 through the postpandemic period. Science, April 2020. Publisher: American Association for the Advancement of Science Section: Report.

[4] Steven A. Frank. Time Delays. In Steven A. Frank, editor, Control Theory Tutorial: Basic Concepts Illustrated by Software Examples, SpringerBriefs in Applied Sciences and Technology. Springer International Publishing, Cham, 2018.

[5] Rifat Sipahi, Silviu-Iulian Niculescu, Chaouki Abdallah, Wim Michiels, and Keqin Gu. Stability and Stabilization of Systems with Time Delay. Control Systems, IEEE, 31, March 2011.

[6] Lukas Richter, Daniela Schmid, and Ernst Stadlober. Methodenbeschreibung für die Schätzung von epidemiologischen Parametern des COVID19 Ausbruchs, Österreich.

[7] Anne Cori, Neil M. Ferguson, Christophe Fraser, and Simon Cauchemez. A New Framework and Software to Estimate Time-Varying Reproduction Numbers During Epidemics. American Journal of Epidemiology, 178(9), November 2013.

[8] Anne Cori. EpiEstim: Estimate Time Varying Reproduction Numbers from Epidemic Curves, 2019. R package version 2.2-1.

[9] Lukas Richter, Daniela Schmid, Ali Chakeri, Sabine Maritschnik, Sabine Pfeiffer, and Ernst Stadlober. Schätzung des seriellen Intervalles von COVID19, Österreich. 2020.

[10] Anne Cori, Neil M. Ferguson, Christophe Fraser, and Simon Cauchemez. A New Framework and Software to Estimate Time-Varying Reproduction Numbers During Epidemics: Web material. American Journal of Epidemiology, 178(9), November 2013.

[11] Sozialministerium. COVID-19: Alle verfügbaren Dateien (gezippt) - data.gv.at. Library Catalog: www.data.gv.at.

[12] Wissen Aktuell: Epidemiologische Parameter des COVID19 Ausbruchs, Österreich, 2020.

[13] Juanjuan Zhang, Maria Litvinova, Wei Wang, Yan Wang, Xiaowei Deng, Xinghui Chen, Mei Li, Wen Zheng, Lan Yi, Xinhua Chen, Qianhui Wu, Yuxia Liang, Xiling Wang, Juan Yang, Kaiyuan Sun, Ira M Longini, M Elizabeth Halloran, Peng Wu, Benjamin J Cowling, Stefano Merler, Cecile Viboud, Alessandro Vespignani, Marco Ajelli, and Hongjie Yu. Evolving epidemiology and transmission dynamics of coronavirus disease 2019 outside Hubei province, China: a descriptive and modelling study. The Lancet Infectious Diseases, April 2020.

[14] R Core Team. R: A Language and Environment for Statistical Computing. R Foundation for Statistical Computing, Vienna, Austria, 2020.

[15] Julie C. Blackwood and Lauren M. Childs. An introduction to compartmental modeling for the budding infectious disease modeler. Letters in Biomathematics, 5(1), December 2018.

[16] Erläuterungen zum Amtlichen Dashboard COVID19, April 2020.

